## Appendices I, II, III for "Knowledge, uptake and intention to use antibiotic post-exposure prophylaxis and meningococcal B vaccine (4CMenB) for gonorrhoea among a large, online community sample of gay, bisexual and other men who have sex with men in the UK"

### Supporting information

Appendix I. Excerpt of RiiSH 2023 survey questions and preambles used for study outcome measures

Appendix II: RiiSH 2023 participant flowchart

Appendix III: Antibiotic regimens ever used as antibiotic post-exposure prophylaxis (PEP) among RiiSH 2023 participants

### Appendix I. Excerpt of RiiSH 2023 survey questions and preambles used for study outcome measures

**Excerpt 1: Questions assessing STI prophylaxis knowledge and use**

**(Preamble)**
There is growing evidence from research studies that taking an **antibiotic** called doxycycline **after having sex** (i.e. doxycycline post-exposure prophylaxis or doxy PEP) can reduce the chances of getting some bacterial STIs (i.e. chlamydia, syphilis and gonorrhoea). Using antibiotics in this way is not currently recommended by clinicians or public health professionals in the UK [click here for information about **doxy PEP** DoxyPEP — QueerHealth] and is different than being prescribed antibiotics to treat an STI that's been diagnosed at a clinic or by testing.

**(Q49.C)**
**Routing: Asked to all participants**
Before taking this survey, had you **heard about** using antibiotics immediately **after sex** to prevent STIs other than HIV (e.g. doxy PEP)?

- No
- Yes
- Don't know

**(Q50. C)**
**Routing: Asked to all participants**
Have you **ever used** antibiotics in this way?

- No
- Yes

**(Q51. C)**
**Routing: Asked to participants who did not report antibiotic use (i.e., Q50C = 'No')**
Have you **ever considered** taking antibiotics in this way?

- No
- Yes

**(Q52. C)**
**Routing: Asked to participants who did report antibiotic use (i.e., Q50C = 'Yes')**
When did you **last use** antibiotics in this way?

- Less than one year ago
- One to two years ago
- More than two years ago

**(Q53. C)**
**Routing: Asked to participants who reported antibiotic use less than one year ago (i.e., Q50C = 'Yes' & i.e., Q52C = 'Less than one year ago')**
Thinking back to the **last** time you used antibiotics in this way, was it:

- Before August 2023
- Since the start of August 2023

**(Q54. C)**
**Routing: Asked to participants who did report antibiotic use (i.e., Q50C = 'Yes')**
Which antibiotic(s) have you used to **prevent** STIs? [tick all that apply]

- Doxycycline (e.g. Vibramycin-D, Efracea, Periostat)
- Azithromycin (e.g. Zithromax)

- Amoxicillin (e.g. Respillin)
- Metronidazole (e.g. Acea, Anabact, Flagyl, Metrogel, Metrosa, Rosiced, Rozex, Vaginyl, Zidoval, Zyomet)
- I don't know
- Other

**Excerpt 2: Questions assessing doxyPEP and Bexsero intention to use**

**(Q55. C)**
**Routing: Asked to all participants**
**Doxycycline PEP** (doxycycline post-exposure prophylaxis or doxy PEP) would involve taking antibiotic tablets within 72 hours **after sex** to try and **prevent getting bacterial STIs**. If this was shown to be safe and effective, **how likely** would you be to take doxy PEP?

- Very unlikely
- Somewhat unlikely
- Somewhat likely
- Very likely
- I don't know

**(Q56. C)**
**Routing: Asked to all participants**
New research evidence suggests that **Bexsero**, a vaccine that protects against a type of meningitis, can lower the risk of getting gonorrhoea by 30%-50%. If this vaccine was available at a sexual health clinic, **how likely** would you consider having it (i.e getting a jab at two different times) to **protect** yourself from gonorrhoea?

- Very unlikely
- Somewhat unlikely
- Somewhat likely
- Very likely
- I don't know

### Appendix II: RiiSH 2023 participant flowchart

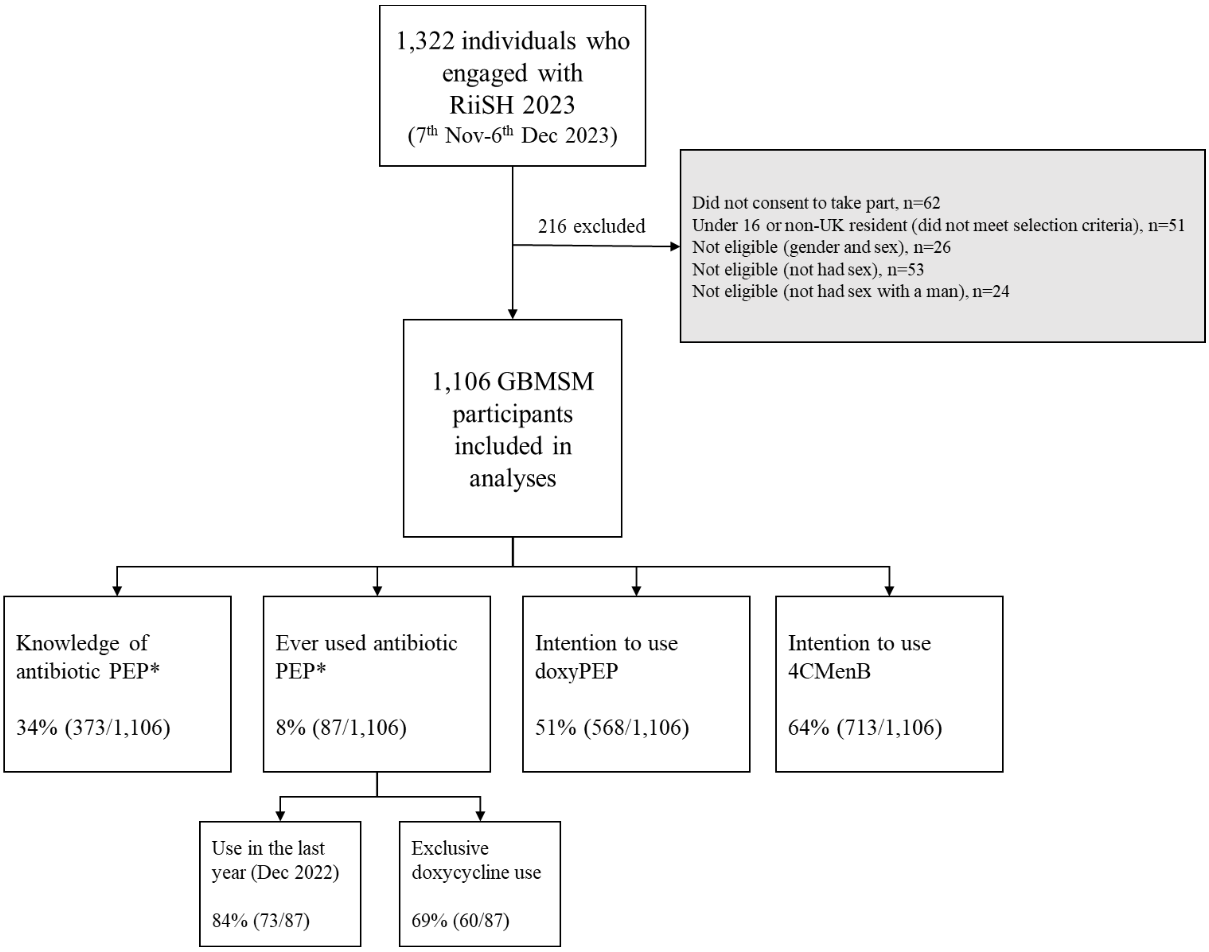

*See Appendix I for survey questions and preambles; report of knowledge and use of antibiotics after sex for STI prevention (i.e., antibiotic PEP). PEP=post-exposure prophylaxis. doxyPEP=doxycycline post-exposure prophylaxis.

### Appendix III: Antibiotic regimens ever used as antibiotic post-exposure prophylaxis (PEP) among RiiSH 2023 participants

| | n (%) |
|---|---|
| RiiSH participants ever reporting antibiotic PEP use* | 87 (100%) |
| Doxycycline only‡ | 60 (69%) |
| Azithromycin only‡ | 2 (2%) |
| Amoxicillin only‡ | 7 (8%) |
| Two or more antibiotics, including doxycycline | 8 (9%) |
| Don't know | 10 (11%) |

*See Appendix I for survey questions and preambles; report of knowledge and use of antibiotics after sex for STI prevention (i.e., antibiotic PEP). ‡ Exclusive use. PEP=post-exposure prophylaxis. doxyPEP=doxycycline post-exposure prophylaxis.
